# Analysis of Respiratory Kinematics: a method to characterize breaths from motion signals

**DOI:** 10.1101/2021.09.06.21263179

**Authors:** William B. Ashe, Sarah E. Innis, Julia N. Shanno, Camille J. Hochheimer, Ronald D. Williams, Sarah J. Ratcliffe, J. Randall Moorman, Shrirang M. Gadrey

## Abstract

**Rationale:** Breathing motion (respiratory kinematics) can be characterized by the interval and depth of each breath, and by magnitude-synchrony relationships between locations. Such characteristics and their breath-by-breath variability might be useful indicators of respiratory health.

**Objectives:** To enable breath-by-breath characterization of respiratory kinematics, we developed a method to detect breaths using motion sensor signals.

**Methods:** In 34 volunteers who underwent maximal exercise testing, we used 8 motion sensors to record upper rib, lower rib and abdominal kinematics at 3 exercise stages (rest, lactate threshold and exhaustion). We recorded volumetric air flow signals using clinical exercise laboratory equipment and synchronized them with kinematic signals. Using instantaneous phase landmarks from the analytic representation of kinematic and flow signals, we identified individual breaths and derived respiratory rate signals at 1Hz. To evaluate the fidelity of kinematics-derived respiratory rate signals, we calculated their cross-correlation with the flow-derived respiratory rate signals. To identify coupling between kinematics and flow, we calculated the Shannon entropy of the relative frequency with which kinematic phase landmarks were distributed over the phase of the flow cycle.

**Measurements and Main Results:** We found good agreement in the kinematics-derived and flow-derived respiratory rate signals, with cross-correlation coefficients as high as 0.94. In some individuals, the kinematics and flow were significantly coupled (Shannon entropy < 2) but the relationship varied within (by exercise stage) and between individuals. The final result was that the phase landmarks from the kinematic signal were uniformly distributed over the phase of the air flow signals (Shannon entropy close to the theoretical maximum of 3.32).

**Conclusions:** The Analysis of Respiratory Kinematics method can yield highly resolved respiratory rate signals by separating individual breaths. This method will facilitate characterization of clinically significant breathing motion patterns on a breath-by-breath basis. The relationship between respiratory kinematics and flow is much more complex than expected, varying between and within individuals.

## 1.0) Introduction

A clinician’s bedside physical examination is never complete without careful characterization of the breathing motion (respiratory kinematics) (1–3). Features of interest include breath intervals, overall depth of breathing and the magnitude-synchrony relationships between key anatomical locations (4). They contain information about the subject’s health which can be useful in wide range of respiratory and non-respiratory illnesses. Tachypnea (short breath interval) and hyperventilation (increased depth of breaths) in the setting of infection can herald the onset of sepsis (5). *Abdominal paradox* (inward abdominal motion, asynchronous with rib cage expansion), in the setting of pneumonia, suggests inspiratory muscle overload and is a risk marker for imminent acute respiratory failure (4,6).

In many clinical scenarios, breath-to-breath variability of motion patterns is also an important feature. Respiratory *alternans* is a sign of inspiratory muscle overload where ribcage-predominant breaths alternate irregularly with abdomen-predominant ones (4,6). Tidal volume variability (shallow breaths alternating with deep ones) precedes hypoventilation (consistently shallow breaths) in opiate-induced respiratory depression and can provide early warnings of an overdose (7–9).

Despite well-established clinical significance, quantitative characterization of respiratory kinematics has proven elusive. Some methods use chest impedance or electrocardiograms to characterize respiratory rates and rhythms (10–12). These can be deployed in intensive care and other telemetry enabled units. Other methods like pneumography, inductance plethysmography, optoelectronic plethysmography, and magnetometry involve obtrusive hardware and/or cumbersome calibration protocols (4,13). They remain limited largely to sleep, exercise and physiology laboratories. In most clinical settings (home, clinics, ambulances and low acuity wards), an experienced clinician’s visual examination remains the sole method to characterize breathing motion patterns.

We have developed the Analysis of Respiratory Kinematics (ARK) system for quantitative characterization of breathing motion in an unobtrusive manner in any clinical setting. Signals are recorded using a network of microelectromechanical system inertial measurement units (MEMS-IMUs) placed on key anatomical landmarks of the chest and abdomen. To pave the way for characterization on a breath-by-breath basis, a method was needed to accurately detect individual breaths using ARK system signals. The aim of this study was to develop a reliable and reproducible method for breath separation that is based on fundamentally sound analytic principles.

## 2.0) Methods

We recruited 34 healthy volunteers for maximal oxygen consumption (VO_2_-Max) testing on a bicycle ergometer in the University of Virginia’s Exercise Physiology Core Laboratory. We placed 8 inertial sensors on key anatomical locations of the chest and abdomen (Figure 1a). Each motion sensor captured 9 streams of signals (3 orthogonal axes each from accelerometers, rate gyroscopes, and magnetometers). To identify breath intervals in this study, we used the linear acceleration signal that was normal to the body surface. We will refer to this signal as the accelerometer signal. To study the relationship of kinematic signals with the air flow cycle, we recorded volumetric flow rate (“flow”) using the laboratory’s standard clinical equipment (Vyaire V-Mask 29-CE; Hans Rudolph metabolic masks).

**Figure 1.**
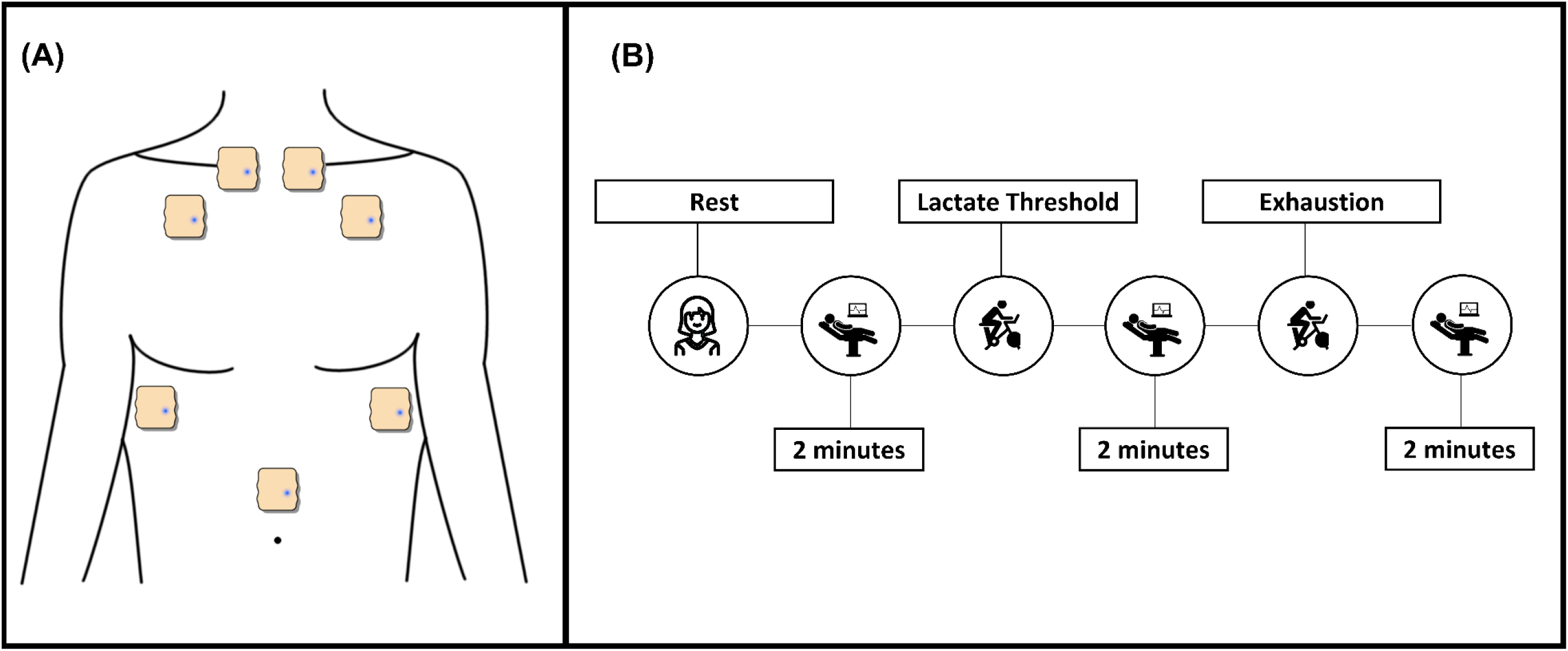
Panel A shows the position of our sensors. The topmost sensors were placed at the insertion of sternocleidomastoid, near the sternoclavicular joint. The second rib sensors were placed in the mid-clavicular line and the eighth rib sensors were placed in the anterior axillary line. The abdominal sensor was placed in the midline abdomen, at that spot above the umbilicus where respiratory motion was most prominently visible to the technician. A sensor was also placed at the base of the neck in the posterior midline (not shown in the figure) to capture non-respiratory motions of the torso. The schematic in Panel B depicts our data acquisition process. We obtained 2-minute recordings of all signals at rest (pre-exercise), lactate threshold, and exhaustion with volunteers in a supine position with 30° head elevation.

We used the Rated Perceived Exertion (RPE) scale to monitor the level of exertion (14), with lactate threshold defined as RPE of 13 (15) and exhaustion as RPE of 20 and/or failure to maintain a pedaling cadence of 60 revolutions per minute. We obtained 2-minute recordings of all signals at rest (pre-exercise), lactate threshold, and exhaustion (Figure 1b). During recording periods, volunteers assumed a supine position with 30° head elevation.

### 2.1) Time synchronization

Each of the eight sensors was connected via USB to a host application running on a computer. Data collection on the sensors was stagger-started over eight consecutive ticks of a common 100Hz timer on the host machine. Once started, each sensor recorded signals at a sampling frequency of 100Hz based on a local timer on the sensor. Data were realigned to a common time frame using the initial host clock start time and the stagger order of the devices. Due to minor inconsistencies between local timers on the sensors, the data were interpolated to identical sampling times across all sensors. For interpolation, data were upsampled to a common 10μS clock by duplicating samples and filtering, then decimated to fall on hundredth-second boundaries.

### 2.2) Filtering

We filtered the flow and accelerometer signals to preserve the respiratory content and to reduce the amplitude of the non-respiratory components. We used a Butterworth bandpass filter (fourth order low pass and sixth order high pass; zero-phase non-causal) with corner frequencies of 0.05Hz and 1Hz. We based this choice on the fact that the frequency of human respirations can reasonably be expected to range between the corresponding rates of 3 and 60 breaths per minute in most circumstances, including after maximal exercise. Since both flow and accelerometer measured the effects of the same underlying cyclical phenomenon – the subject’s respiration – we used the same filter specifications for both signals.

### 2.3) Identifying breath intervals using the analytic representation of filtered signals

From the filtered signals (both flow and acceleration), we generated the corresponding analytic representations. For any real valued signal *u*(*t*), its analytic representation *u*_*a*_(*t*) is defined as:

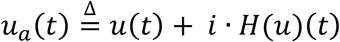

Here, *H*(*u*)(*t*) is the Hilbert transform of *u*(*t*), which shifts the phase of its components by 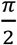 radians for negative frequencies and by 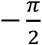 radians for positive frequencies (16). In the analytic representation, the Hilbert transform is plotted on the imaginary y-axis [*i* · *H*(*u*)(*t*)] as a function of the untransformed signal in the real x-axis of a complex plane. Multiplying by *i* shifts all phases by an additional 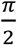 radians, restoring the phase of positive frequency components while negating the negative frequency components when added to the original, real signal.

The angle made between points on the analytic signal and the positive real axis, with the origin as the vertex, represents the time-varying phase of the wave. Since the analytic representation contains only positive frequencies, the resulting instantaneous phase angle (ϕ) of the wave monotonically increases to match the progression of the complex signal around the origin. For quasi-cyclic processes with repetitive but not strictly periodic behavior, this instantaneous phase angle can be used to reliably detect consistent landmarks, where a given phase closely tracks the same point on the original signal (e.g. a specific peak or zero-crossing) across different cycles. We used this property to identify breath intervals on flow and accelerometer signals as the intervals between successive occurrences of a particular phase angle (Figure 2).

**Figure 2.**
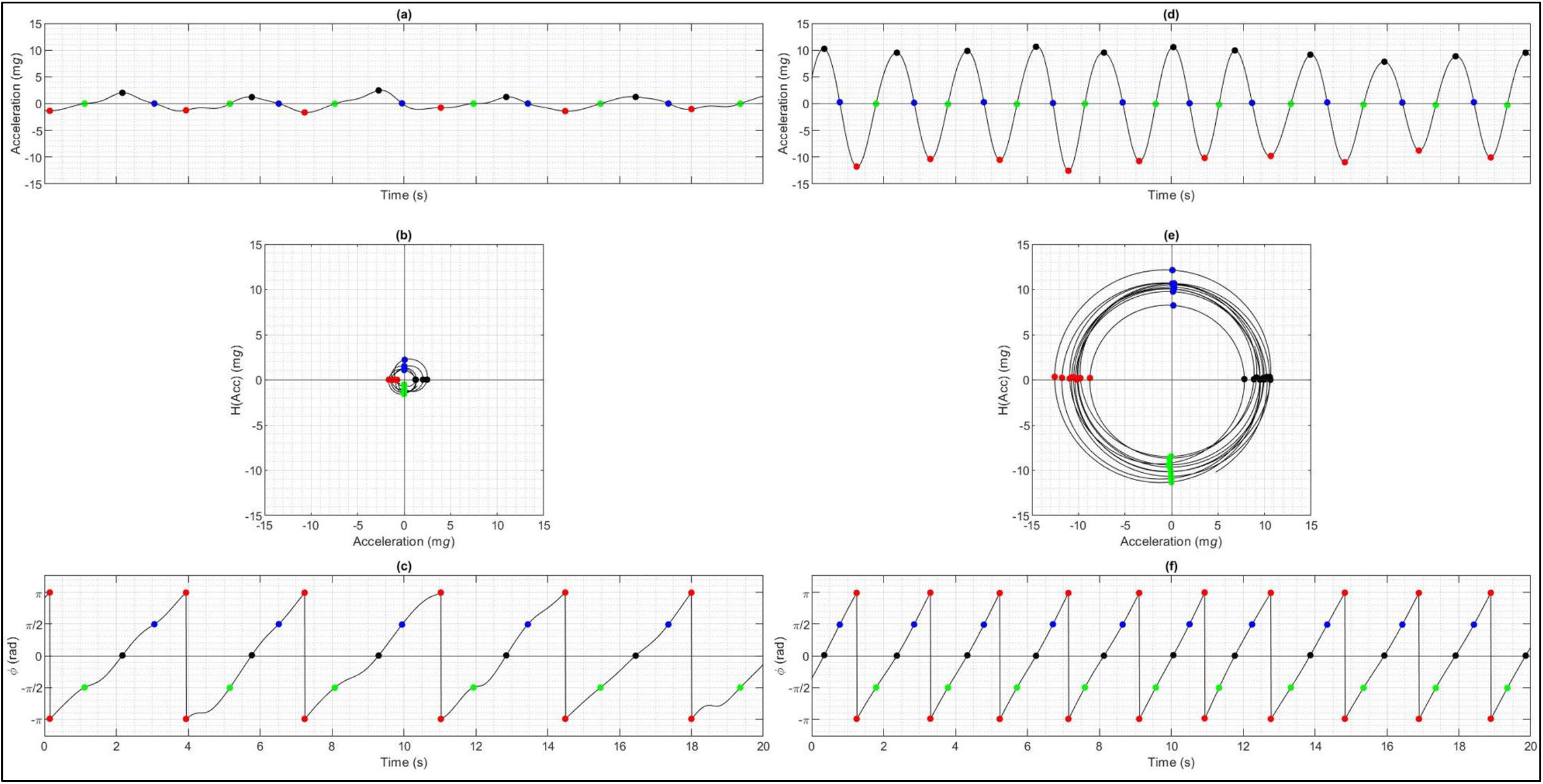
Selecting landmark points using the phase angle from its analytic representation. Panels A, B and C show resting signals and panels D, E, F show signals at exhaustion from the same subject. Panels A and D show the acceleration signal in time domain; panels B and E show the corresponding analytic representations. In an analytic representation of a signal, the Hilbert transform of the signal is plotted, on an imaginary y-axis of a complex plane, as a function of the untransformed signal. In this complex plane, the phase angle for any number is defined as angle between the positive real axis and the line joining the origin and that number. Panel C shows a time domain plot of the phase angle. Across all panels, the points with phase angles of 0, 0.5π, π and -0.5π radians are colored black, blue, red, and green respectively.

### 2.4) Evaluating fidelity of accelerometer-derived breath intervals

We derived respiratory rate signals from accelerometer and flow signals by adapting the method that was originally proposed by Berger and coworkers to derive heart rate signals from electrocardiograms (17). We used intervals between instantaneous phase landmarks on the accelerometer and flow signals in place of R-R intervals on electrocardiograms (Figure 4a). In this study, we chose the phase angle of π radians as our landmark for both flow and accelerometer signals, and we resampled the respiratory rate at 1Hz. To evaluate the fidelity of accelerometer-derived breath intervals, we calculated cross-correlation between accelerometer-derived and flow-derived respiratory rate time series.

### 2.5) Relating the phase of flow and accelerometer signals

We calculated the relative frequency with which kinematic phase landmarks were distributed over the phase of the air flow cycle. We split the air flow cycle into 10 bins (bin width of 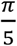 radians). Similar methods have been used in the past to create cardiorespiratory synchrograms (18). To quantify synchronization, we calculated the Shannon Entropy of the histograms:

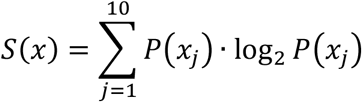

Here *P*(*x*_*j*_) is the probability (relative frequency) of a kinematic phase landmark occurring in the *j*^th^ bin of air flow phase. The utility of this measure in quantifying cardiopulmonary coupling has been described (19). In our application, the Shannon entropy could range between a maximum of 3.32 bits (log_2_10) signifying uniform distribution across 10 bins, and a minimum of 0 bits signifying localization to a single bin (Figure 4b).

We used MATLAB version 2020b for signal processing (20). We used REDCap electronic data capture tools hosted at the University of Virginia (21,22).

## 3.0) Results

Of the 34 volunteers, five had no usable data and seven had only partial data. The final sample consisted of 22 healthy volunteers. The reasons for missing data were related to unanticipated hardware and software malfunction in our locally produced prototype. Our subjects, a third of whom were women, had a mean age of 47 (range: 21-71), a mean BMI of 25 (range: 21-33) and a mean VO_2_-max of 42 ml/kg/min (range: 15-60).

### 3.1) Illustrative examples of results

Figure 2 shows examples of acceleration signals (Panels A and D), their analytic representations (Panels B and E), and time domain plots of instantaneous phase angles (Panels C and F). The color-coded dots demonstrate the property that each occurrence of a particular phase angle is separated from the last by one respiratory cycle. We can also note that points of maxima and minima in the untransformed signal correspond with the phase angles of 0 and π on the analytic representation. Similarly, zero crossings on the untransformed signal correspond with phase angles of 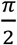 and 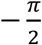 on the analytic representation. For the flow signals, these points are clinically significant and represent onset of inhalation 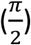, onset of exhalation 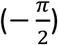, peak inspiratory flow rate (0), and peak expiratory flow rate (π) respectively.

Figure 3 shows landmarks (phase angle of π) plotted as vertical lines on the signals from which they were derived. In all panels that capture a meaningful degree of respiratory motion, the interval between these landmarks was defined as the accelerometer-derived breath interval. These intervals were used to (a) derive a respiratory rate signal (Figure 4a) and (b) identify coupling between kinematics and flow (Figure 4b).

**Figure 3.**
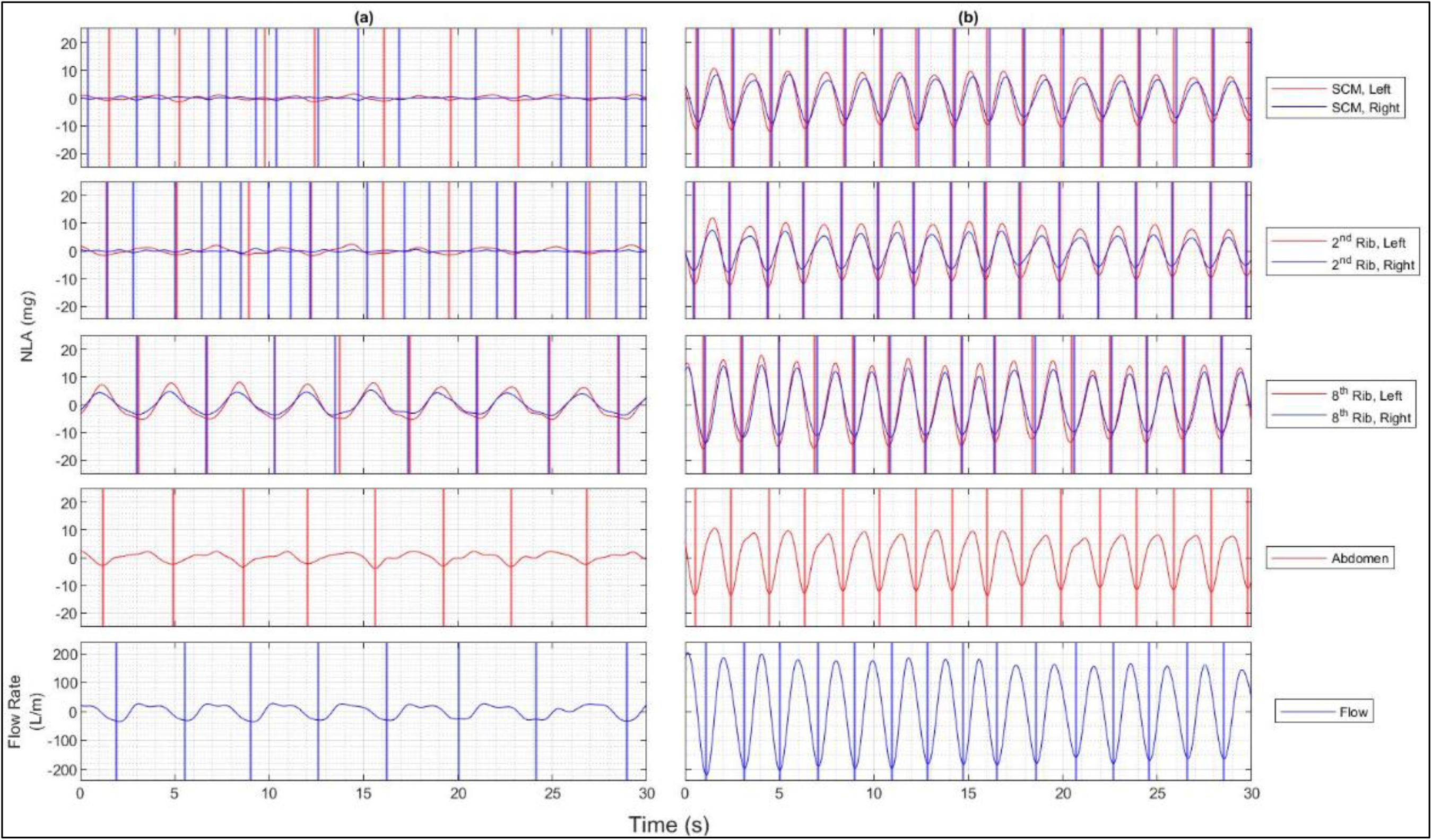
Visualizing respiratory kinematics: This figure shows 30 second strips of accelerometer signals obtained at rest (left) and after maximal exercise (right) by the same individual. The top four panels are organized by kinematic sensor location: sternocleidomastoid insertion (SCM), 2^nd^ rib, 8^th^ ribs & midline abdomen respectively. For bilateral sensors, color encodes laterality - blue is right and red is left. The bottom panel shows air flow signals recorded using exercise laboratory equipment. The vertical lines mark the location of landmark points selected using a phase angle of π radians. The intervals between these landmarks decrease sharply, reflecting tachypnea after exertion. Additionally, significant changes can be noted in magnitude and synchrony of motion at various sensor locations. Most prominently, thorax predominant breathing at rest changes to a mixed thoraco-abdominal breathing at exhaustion (comparing signals at 8^th^ rib and abdomen). Upper thoracic motion (SCM and 2^nd^ rib), which likely reflects accessory respiratory muscle recruitment, rises from being negligible to being comparable to lower thoracic signals at exhaustion

**Figure 4.**
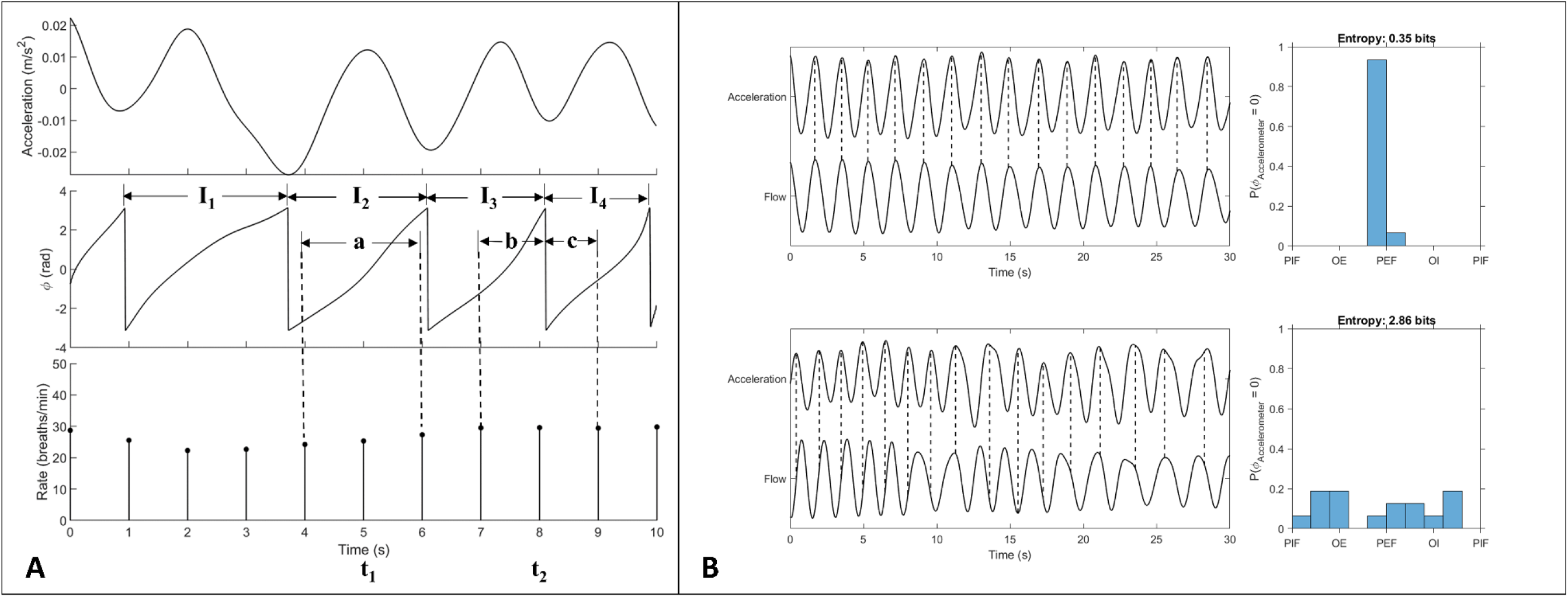
(A) **<u>Respiratory rate sampling:</u>** The top two curves show a segment of the acceleration signal and its instantaneous phase. The respiratory rate samples derived from these intervals are shown at the bottom. First, breath intervals (labelled as I_1_ to I_4_) are determined using consecutive occurrences of a phase angle (π radians, in this case). Next, a sampling rate for the respiratory rate (f_r_) signal is chosen as desired (1 Hz in our case), without regard to mean respiratory rate or sampling frequency of the acceleration signal. For each sampling point, we count the number of breath intervals (n_i_), including fractions, that occur in the time window extending from the previous sample to the next. For example, at time 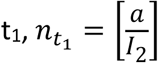 and at time 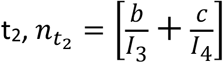. The respiratory rate (r_i_) at each sampling point is calculated as 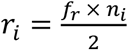. **(B) <u>Kinematics-flow coupling:</u>** Here, we illustrate our method to measure coupling between signals using one example each of strong (top) and weak (bottom) coupling. When coupling is strong, landmarks from the phase of the acceleration signals (phase of 0 radians, in this case) are strongly localized to a particular portion of the phase of the flow cycle (mid-exhalation, in this case), resulting in low entropy (0.35 bits in this case). When coupling is weak, landmarks are uniformly distributed over the phase of the flow phase, resulting in high entropy (2.86 bits in this case). [PIF: Peak Inspiratory Flow; OE: Onset of Exhalation; PEF: Peak Expiratory Flow; OI: Onset of Inhalation; ϕ = Phase angle]

Figure 5 shows four examples where flow-derived respiratory rate (solid black line) is plotted alongside accelerometer-derived respiratory rate signal. Panel A is a resting series with a very stable respiratory rate. The other panels show diverse patterns of recovery from exhaustion – a smooth downward trend in Panel B; a cyclical fluctuation superimposed on a downward trend in Panel C; and a slow recovery interrupted by a sharp transient deceleration in Panel D. Each of these patterns were reproduced with high fidelity in the accelerometer-derived respiratory rate signal.

**Figure 5:**
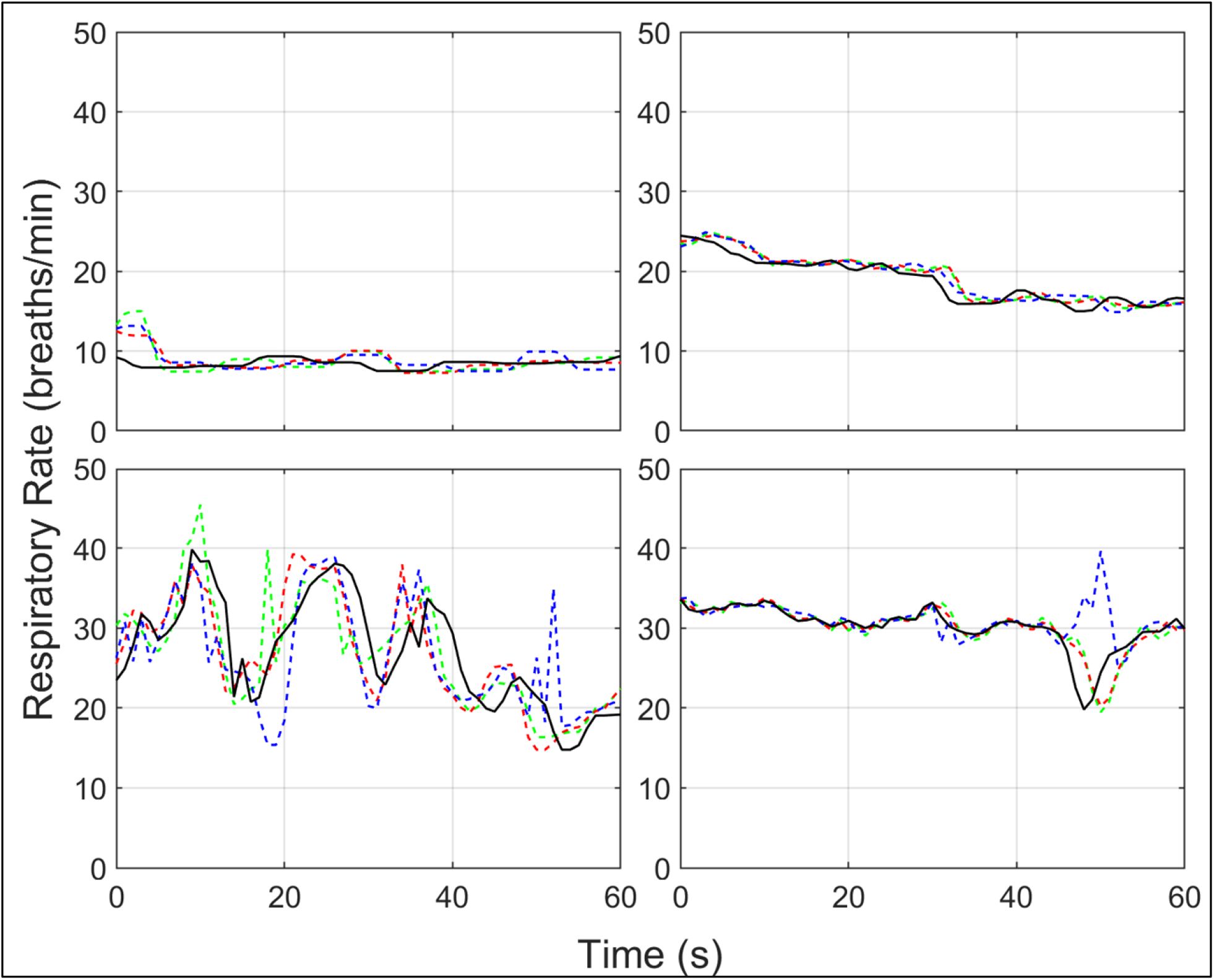
Respiratory rate signals. This figure highlights the diversity of respiratory rate signals that we observed in our small sample. The rate signals plotted in the solid black line were derived from air flow signals whereas dotted lines are rate signals sampled from 8^th^ rib (red and green) and abdominal (blue) acceleration signals. Panel A is a resting series with a very stable respiratory rate. Panels B, C and D are all series obtained at exhaustion and show a variable recovery pattern. In panel B, there is a smooth recovery in respiratory rate. Panel C shows a cyclical fluctuation of respiratory rate in addition to a downward trend. In Panel D, we see very gradual recovery that is interrupted with a transient slow-down. In each of these instances, respiratory rate signals were reproduced with high fidelity using the ARK method.

Figure 6 shows examples of the complexity and diversity that we observed in the coupling between kinematics and flow. Panel A shows the results for an individual where the kinematics and flow were significantly coupled in all states of exercise – Shannon entropies of 1.87, 1.55 and 1.24. Interestingly, the point of localization varied by exercise stage - occurring in mid exhalation, at peak inspiratory flow, and at onset of inhalation during rest, lactate threshold and maximal exercise respectively. The overall localization was poor (Shannon entropy of 2.83) when all breaths were considered without regard to exercise state. In some other individuals, we found no localization even within a stage (Panel B).

**Figure 6:**
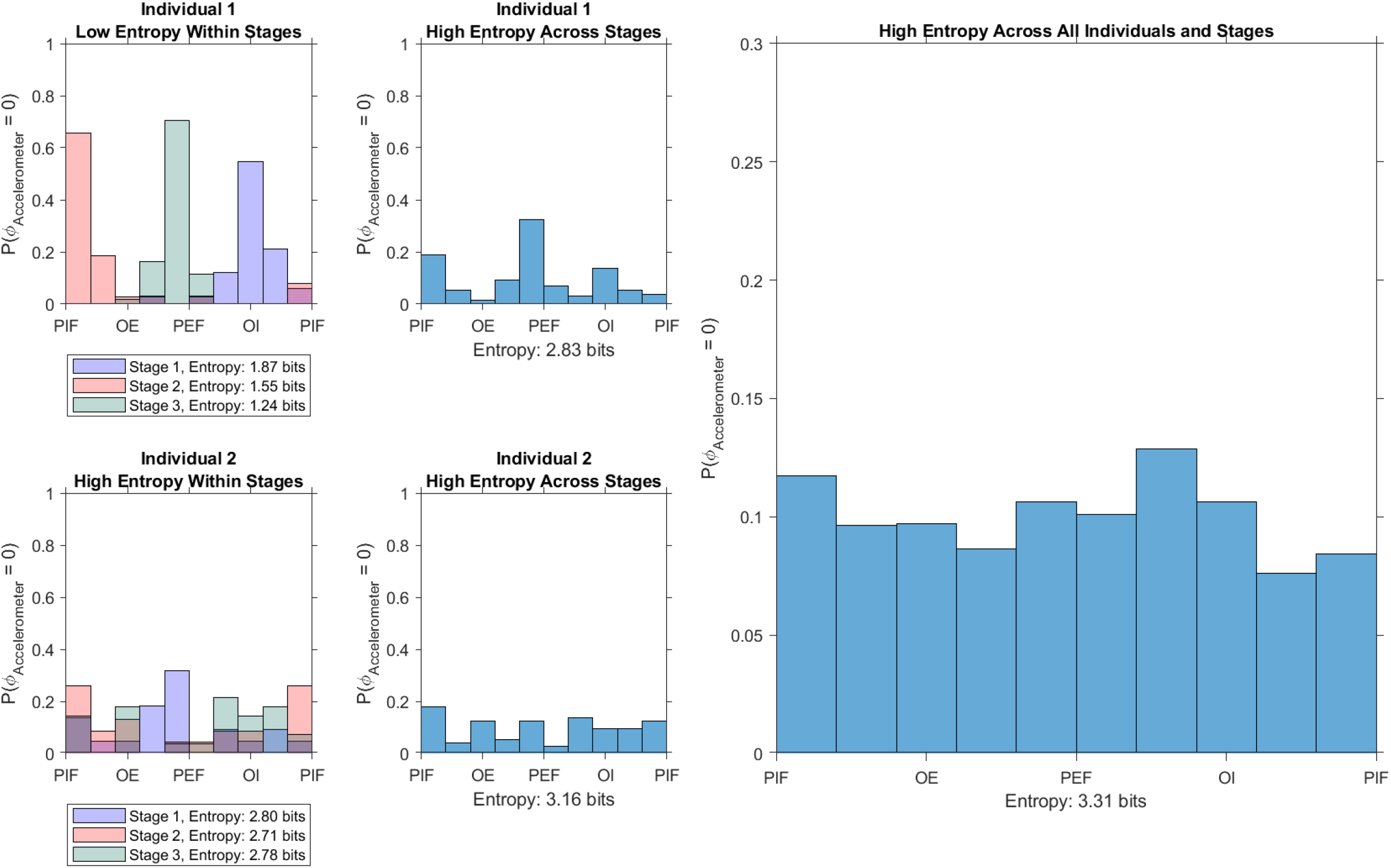
Kinematics-Flow Coupling: This figure illustrates the observation that kinematics and flow have a complex relationship which varies between and within individuals. Panel A shows findings from an individual whose kinematics were strongly coupled with flow within a stage. In each stage, however, the kinematic phase landmarks localized to different areas of the air flow phase. As a result, the overall Shannon entropy for that individual was higher (2.83) than their stage-specific Shannon Entropy (1.87, 1.55, 1.24). Panel B shows findings from an individual whose kinematics weakly coupled with flow even within a stage. Across individuals and exercises stages, therefore, the overall Shannon entropy (3.31) was very close to the maximal possible value of 3.32 (panel C). This figure displays results obtained from a lower rib sensor. Similar results were noted at all sensor locations. [PIF: Peak Inspiratory Flow; OE: Onset of Exhalation; PEF: Peak Expiratory Flow; OI: Onset of Inhalation; ϕ = Phase angle]

### 3.2) Summary Results

We found a strong relationship between respiratory rate time series derived from accelerometer and flow signals. In light of the variability we observed in the kinematic-flow cycle synchronicity, we quantified the degree of agreement using the maximal cross-correlation value without regard for lag. We observed cross-correlation coefficients as high as 0.94. Of note, the strength of this relationship was preserved despite trends, cyclical fluctuations or transient disturbances in true respiratory rate (Figure 5).

When analyzed across individuals and exercise states, landmarks from the phase of kinematic signals were uniformly distributed across the phase of the air flow cycle (Figure 6). The average Shannon entropy across the lower rib and abdominal sensors was 3.31, which is comparable to the entropy limit of 3.32 for a completely uniform distribution.

## Discussion

The clinical significance of breathing motion patterns is well established, but there are no unobtrusive methods to quantitatively characterize such patterns at the bedside. Clinicians must rely on visual inspection to assess breathing motion, and qualitatively describe their findings in notes. This is problematic for several reasons. First, there is considerable inaccuracy in manual recordings of even the simplest kinematic feature, the respiratory rate (23,24). For more complex features, even highly trained physicians achieved only moderate inter-rater reliability (25). Less experienced clinicians may simply miss red-flag signs or fail to recognize the risk they portend (26). Second, attempts to standardize terminology in narrative descriptions have not achieved wide adoption (27). Such an approach would require large scale continuous training of staff at every level of care. Finally, only a small proportion of variance in respiratory kinematic features is explained by conventionally monitored vital signs (28). Timely detection of respiratory kinematic aberrations, therefore, depends on frequent bedside assessments by experienced clinicians. This is not possible in most settings due to resource and/or staffing constraints. Patient isolation requirements in illnesses like COVID-19 further reduce the frequency of bedside assessment (29–31). Visualizations like the ones in Figure 3 or 6 can facilitate in-depth inspection of kinematic features by clinicians. They can be reviewed remotely and stored as a reference to ascertain the trajectory of respiratory kinematics over time. Quantitative characterization of the signals will yield novel features which may prove useful in predictive modelling of respiratory failure. We foresee considerable clinical utility in such remote, quantitative assessments of respiratory effort.

We found that the relationship between kinematic and air flow cycles was rich with complexity – varying between and within individuals. For practical breath-detection applications, this finding has an important implication – no landmark point is superior to any other in terms of optimizing alignment between the kinematic and air flow cycles. We selected the phase angle of π radians to identify each nadir in the accelerometer signals. Any other landmark (like the phase angle of 0 radians to identify each peak of the accelerometer signal) might be selected.

We demonstrated that the ARK signals can yield highly resolved respiratory rate time series from kinematic signals. This finding paves way for two types of analyses in the future. First, a large number of well-established mathematical operations can be used to characterize respiratory rate time series in any clinical setting (32). The regular sampling on a real-time axis also allows for meaningful analysis in the frequency domain. Second, well-defined breath intervals will allow breath-by-breath characterization of signal features, *i*.*e*., comparing magnitude-synchrony relationships between different sensor locations within a single kinematic cycle. This is important to quantify patterns like respiratory *alternans* or opiate-induced ataxic breathing, where the breathing pattern varies from one breath to the next. Our future work will build on this breath detection method to describe the clinical significance of a large and diverse set of metrics. The overarching vision is to create a set of novel respiratory vital signs that can be conveniently measured in any setting and improve medical decision making in common clinical scenarios.

A strength of this work is that it is grounded in fundamentally sound analytic methods like the Hilbert transform, the analytic representation of a signal, Shannon entropy, and Berger’s interpolation algorithm. We do not rely on any arbitrary assumptions, decisions, thresholds or models. This enhances the reliability, reproducibility of our method and its generalizability to other signals that record respiratory motion (impedance, inductance plethysmography etc.). Another strength is the unique data set. To our knowledge, this is the first instance where multi-focal kinematic signals are compared to synchronously-recorded volumetric air flow signals in varying states of respiratory physiology. This is, therefore, among the first descriptions of the enormous variability encountered in the flow-kinematics relationship.

The small size and inclusion of only healthy individuals are limitations and further development in larger and more clinically diverse samples will be needed. The results at this stage serve as demonstration of feasibility of our breath detection method and lays the groundwork for detailed breath characterization in the future.

## Conclusion

We describe a reliable and reproducible method to detect individual breath cycles from respiratory kinematic signals. Despite a complex relationship between respiratory kinematics and air flow, our method resulted in highly resolved respiratory rate time series. This will facilitate quantitative characterization of clinically significant breathing motion patterns in any care setting.

## Data Availability

We have not made data publicly available.

## Acknowledgements

We thank David Chen, director of UVA’s Coulter Translational Research Partnership, for his role in establishing and supporting this multidisciplinary collaboration. We thank Sharon Krueger, program director of the Ivy Foundation for her advice and feedback. We thank UVA’s Division of General, Geriatric, Palliative and Hospital Medicine, UVA’s Center for Engineering in Medicine and the Ivy Foundation for the seed funding which made this project possible. We thank Dr. Arthur L. Weltman, Director of UVA’s Exercise Physiology Core Laboratory (EPCL), for generously lending his domain expertise to optimize our study design. We thank EPCL’s manager, Lisa Farr, and her entire team (Olivia Hazelrigg, Nicole Huebner, Courtney Paxton and Jackie Dosik) for their enthusiastic support of our work. We thank Melissa Goldman, Fabrication Lab manager at the UVA School of Architecture, and Trevor Kemp, assistant manager, for their advice and assistance throughout prototype design and construction.

## Footnotes

None

## Tables

None

